# Reporting of Qualitative Research Using Large Language Models (COREQ+LLM): A Delphi-based Extension of the COREQ Reporting Guideline

**DOI:** 10.64898/2026.10.01.26364479

**Authors:** Leonard Fehring, Julian Frings, Paul Rust, Christian Kempny, Petra Thürmann, Emre Tamerler, Sven Meister

## Abstract

**Background:** Qualitative research provides insights into human behavior, perspectives, and lived experience. While existing standards like the Consolidated Criteria for Reporting Qualitative Research (COREQ) have improved transparency and rigor, the integration of large language models (LLMs) into qualitative research introduces new challenges for not covered by current guidelines. LLMs transform analytic processes and blur the line between human interpretation and algorithmic text generation, raising concerns about transparency, authorship, and epistemic trust. The here presented COREQ+LLM guideline extends COREQ to establish comprehensive reporting standards for LLM use in qualitative research.

**Methods:** The study followed the EQUATOR Network’s guideline development framework in two phases: (1) a systematic scoping review to identify best practices for reporting LLM use in qualitative research, and (2) a Delphi consensus process with international experts in qualitative research, AI, medicine, ethics, and journal editing to develop a comprehensive reporting checklist.

**Findings:** The scoping review identified 149 studies relevant for full-text review. From this literature, 29 preliminary reporting items were extracted. In the first Delphi round, international experts (N=141) assessed the items’ relevance. 20 items were accepted for inclusion in the guideline (>75% agreement); however, four required reevaluation in the second round, five underwent wording revisions, and six new items were proposed. In the second Delphi round (N=115; retention rate 82%), one was excluded, while four items still lacked consensus. Remaining items were discussed in a virtual consensus conference with nine representatives of the expert panel, resulting in the inclusion of two and the exclusion of two others. The final checklist comprises 31 items grouped into seven categories, serving as an extension of the original COREQ checklist.

**Interpretation:** COREQ+LLM offers structured, consensus-based guidance to enhance transparency, reproducibility, and methodological rigor in qualitative research using LLMs. Its adoption will support researchers, reviewers, and editors in maintaining trust and scientific integrity.

## Introduction

Qualitative research plays a crucial role in health sciences by offering detailed understanding of human behavior, perceptions, and experiences.^1-3^ It relies on non-quantitative approaches to explore social and psychological phenomena and to develop new perspectives.^1 4^ Common approaches are interviews, focus groups, participant observation, and document or artifact analysis, combined with analytic techniques such as coding, thematic analysis, and content analysis.^5 6^

The integration of large language models (LLMs) into qualitative research is rapidly expanding. LLMs are artificial intelligence (AI) systems trained on extensive text datasets to understand and generate human-like language using deep learning, particularly transformer architectures. They perform diverse language tasks. In qualitative research, they are increasingly used to design interview guides, transcribe and translate data, support coding and thematic analysis, and even interact with qualitative datasets.^7-14^ LLM functionality is also being integrated into a growing range of commercial qualitative analysis tools.

The rise of LLMs represents a methodological turning point in qualitative inquiry. Unlike earlier digital tools limited to data management, LLMs can generate, interpret, and transform textual data, thereby challenging conventional boundaries between human interpretation and algorithmic inference. This development forces a reconsideration of how interpretive authority is negotiated between humans and LLMs, striking at the epistemological core of qualitative research.

Moreover, LLMs raise methodological challenges, including non-deterministic outputs, bias propagation,^15 16^ and lack of interpretability. They generate text by predicting the most likely next token based on patterns learned from training data, making their outputs inherently probabilistic rather than deterministic. This means that even when given the same instruction (prompt), they can produce different answers.^17^ In scientific contexts, this probabilistic behavior is challenging because it can lead to unreliable or unverifiable results, complicating verification and reproducibility. Furthermore, LLMs may replicate or amplify biases inherent in their training data and can confidently produce factually incorrect statements, also known as ‘hallucination’.^18^ These characteristics pose risks to the reliability and credibility of qualitative research findings.

Without explicit reporting, the use of LLMs in qualitative analysis risks remaining invisible, potentially undermining the interpretive credibility of findings and the ethical accountability of researchers. Studies may appear human-led while in fact relying substantially on algorithmic synthesis. Transparent disclosure is therefore not merely a matter of methodological completeness but of maintaining public and scholarly trust in qualitative research.

Current reporting guidelines provide limited direction for addressing these challenges.

Developed in 2007, the Consolidated Criteria for Reporting Qualitative Research (COREQ) checklist is a guideline comprising 32 items for reporting key elements of qualitative research (such as the research team, methods, context, findings, analysis, and interpretations).^19^ Its broad adoption across journals and disciplines attests to its effectiveness in promoting consistency and rigor in qualitative reporting, making it the most authoritative framework currently available for qualitative reporting. However, while COREQ includes one item concerning software use (item 27), it does not capture critical aspects of LLM integration such as model selection, configuration, prompting strategies, bias mitigation, or validation.

Existing guidelines for reporting AI use in scientific research (i.e. TRIPOD-LLM, CONSORT-AI, and SPIRIT-AI ^20-22^) focus primarily on the development and evaluation of AI systems in prediction modeling and clinical trials. None explicitly engage with the distinct methodological, ethical, and epistemological challenges posed by the use of LLMs in qualitative research.

The scientific community has highlighted this absence of normative frameworks and transparency in the use of LLMs for qualitative analysis.^23 24^ Findings from a workshop with 40 researchers indicate that LLMs are already integrated into research practices, while standards for evaluating validity and rigor remain undeveloped.^25^

The objective of the present study is to address this gap by developing a reporting checklist for the use of LLMs in qualitative research. Given COREQ’s established credibility and widespread adoption, it serves as the foundation for this initiative. The proposed COREQ+LLM extension aims to provide an evidence-based, consensus-driven framework to ensure rigorous and transparent reporting of LLM use in qualitative studies. Beyond methodological standardization, COREQ+LLM seeks to safeguard interpretive integrity, clarify authorship boundaries between humans and AI systems, and promote responsible innovation in digital qualitative methods. We examine how LLMs are currently reported as qualitative research tools in peer-reviewed studies to generate an initial list of potential reporting items, which is further refined through structured expert consultation via a Delphi consensus process.

## Methods

### Study Design

The development of this reporting guideline followed a multiphase mixed-methods design comprising a scoping review, a two-round classic Delphi survey and a final consensus conference. The detailed methodological framework was previously published as a research protocol^26^ and inspired by other EQUATOR guidelines. The project was prospectively registered with the EQUATOR Network as a guideline under development and with the Open Science Framework for the scoping review component (registration DOI: 10.17605/OSF.IO/BK42Y). Ethical approval was obtained from the Ethics Committee of Witten/Herdecke University (reference S-165/2025). An overview of the methodological approach is depicted in figure 1.

**Figure 1.**
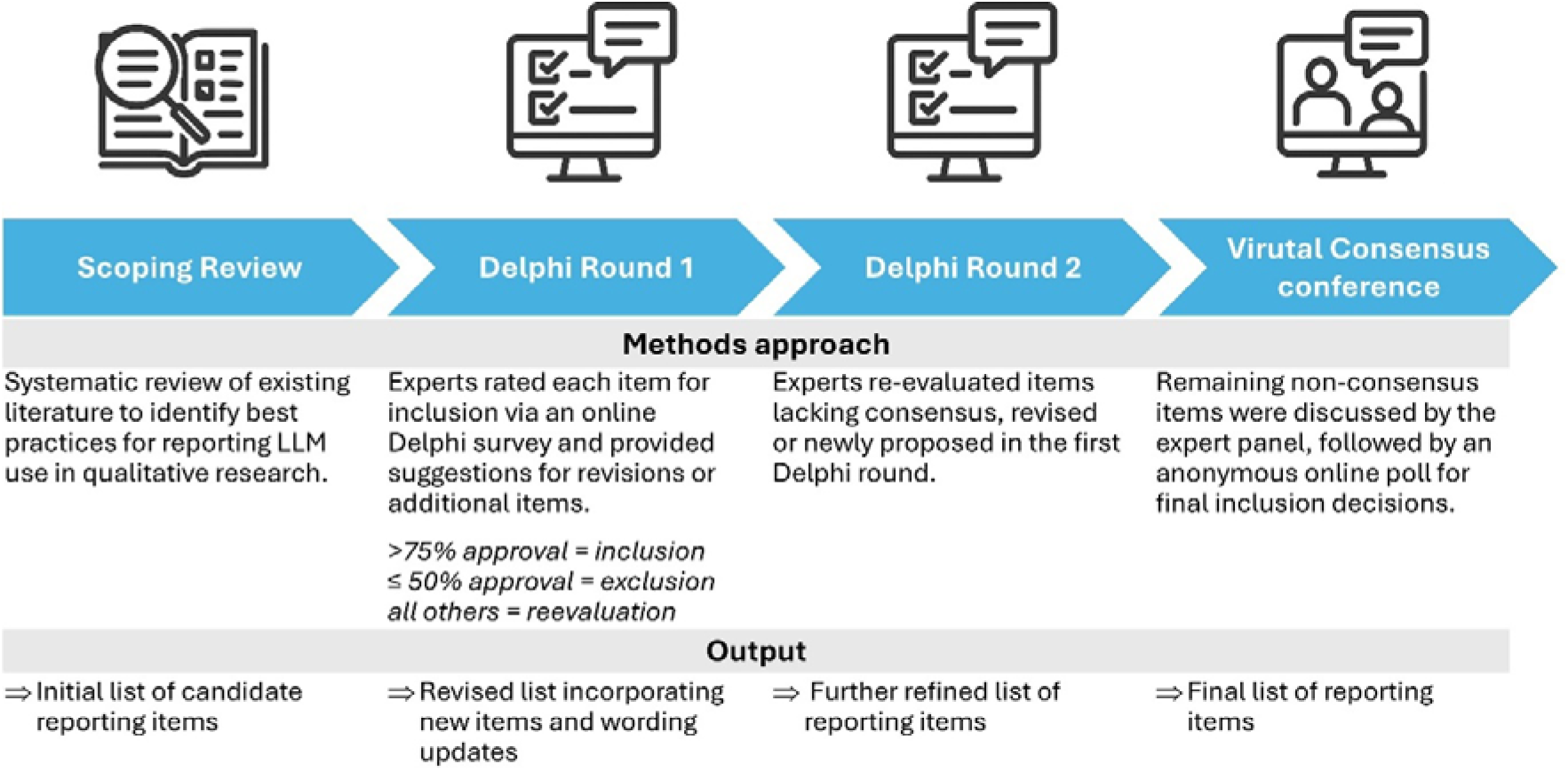
Visualization of the methodological approach.

In this article, we report the methods and results of the Delphi survey and the consensus conference. The scoping review is summarized briefly, with full methods and findings to be published separately.

### Phase 1: Scoping Review

The PRISMA-ScR scoping review examined how the use of LLMs in qualitative research has been reported in published literature. Eligible studies included English-language, peer-reviewed articles published between January 1^st^, 2020, and May 31^st^, 2025, that used LLMs as tools in conducting qualitative research (e.g., for interviewing, transcription, thematic coding, data interpretation, or content synthesis). Studies were excluded if LLMs were used solely for manuscript editing, evaluated as the subject of performance testing rather than as research tools, or if the article lacked an empirical qualitative component (e.g., commentaries, viewpoints, or editorials).

Searches were conducted in June 2025 across PubMed, Scopus, APA PsycINFO, CINAHL, and Business Source Premier. Search strategies combined controlled vocabulary (MeSH terms) and free-text keywords related to “large language models,” “ChatGPT,” “generative AI,” and “qualitative research.”

After duplicate removal, 4,201 unique records were screened independently by three reviewers (PR, JF, CK) in a two-stage process (title/abstract and full-text screening). Discrepancies were resolved through discussion with a fourth reviewer (LF). The relevant articles were screened for details of LLM use (e.g., model type such as GPT-4, Claude, or Gemini), prompting and configuration strategies, and the level of reporting transparency, including bias, ethical considerations, and reproducibility statements. From this information we developed an initial list of candidate items that are suitable to extend the existing COREQ checklist. COREQ was selected as the foundation because it was specifically designed for interview- and focus group–based studies, in which the advantages of LLM-assisted analysis are particularly pertinent. Unlike broader or less systematic frameworks, COREQ provides a structured approach that ensures methodological transparency and consistency, making it the most widely recognized and authoritative standard for qualitative reporting.

### Phase 2: Delphi Consensus Process

To establish expert consensus on which candidate items to include in the final COREQ+LLM checklist, we conducted a two-round online Delphi process between July and October 2025 using LimeSurvey, hosted on a secure university server. The process followed the DELPHISTAR^27^ and EQUATOR Network^28^ recommendations for designing and reporting consensus studies. Supplementary material 1 presents the DELPHISTAR table.

### Participant Recruitment

We identified potential experts through literature searches, professional networks, and snowball sampling and invited them via their publicly available email addresses. In addition, we contacted certified instructors from major qualitative research software providers, including AILyze (https://www.ailyze.com), Lumivero (https://lumivero.com), MAXQDA (https://www.maxqda.com), and ATLAS.ti (https://atlasti.com). Eligible participants demonstrated expertise in qualitative methodology, guideline development, AI ethics, or research involving LLMs.

### Procedure and Analysis

The Delphi study was designed and led by an interdisciplinary team with representatives from medicine, psychology, ethic committees, qualitative research and healthcare informatics. No consulting was received. The first Delphi round was conducted from 22 August to 12 September 2025 (3 weeks), and the second Delphi round from 24 September to 9 October 2025 (15 days).

In each Delphi round, participants rated every candidate item on a 5-point Likert scale (1 = definitely exclude, 2 = rather exclude, 3 = neutral, 4 = rather include, 5 = definitely include). We defined consensus for inclusion a priori as >75% of respondents rating an item as 4 or 5 (=agreement). Items with ≤50% agreement were excluded, while those with >50% and ≤75% agreement were revised and re-evaluated in the following round.

Participants could provide free-text feedback in each round to suggest modifications to item wording or propose additional items. Two researchers (JF, PR) conducted a thematic analysis of the free-text comments in round one following a rapid and pragmatic approach to refine existing items and identify new candidates for round 2. The process involved familiarizing themselves with the responses, coding repeated ideas, and grouping them into themes to refine existing items and identify new candidates for round two. This rapid thematic analysis was adapted from Braun & Clarke’s framework to suit the brief, exploratory nature of the survey comments.^3^ All newly proposed or revised items were included in the second-round survey for re-evaluation. For items without consensus in round 1 approval rates where indicated in round 2. After round 2 two other researchers (LF, ET) conducted the thematic analysis of free text comments.

Items that did not reach consensus after the second Delphi round were carried forward to the Virtual Consensus Conference for final discussion and voting.

### Virtual Consensus Conference

We invited all participants from the second Delphi round to attend the virtual consensus conference, held on October 16, 2025, via the Zoom platform. During the meeting, the remaining items were presented together with the results from Delphi rounds 1 and 2. Participants discussed the potential inclusion until thematic saturation was reached.

Following discussion, participants voted anonymously using Zoom’s built-in polling feature. Items that achieved >75% agreement were included in the final COREQ+LLM checklist. We recorded and transcribed the meeting to ensure transparency and traceability of the decision-making process. Two researchers (LF and ET) systematically analyzed the transcripts to document the key arguments and rationale underlying each consensus decision.

## Results

### Scoping Review

The search strategy of the scoping review identified 5,049 records. After removing duplicates, 4,201 studies (83.2%) progressed to title and abstract screening. Title and abstract screening yielded 149 studies that used LLMs in qualitative health research, which were subsequently included for full-text review.

Most studies described general tasks (e.g., coding assistance or theme generation) performed by the LLM but lacked detail about prompting strategies, model names and versions, model parameters, validation procedures, and human oversight mechanisms. Only a minority explicitly discussed model limitations, bias mitigation, or hallucination management.

The PRISMA-ScR scoping review (currently under peer review and not published yet) yielded, 29 initial candidate items. These items were organized by the research team in an iterative process under seven subheadings: 1) Research team and reflexivity, 2) Study design and theoretical framework, 3) Model and access, 4) Prompting, 5) Context use and session management, 6) Data analysis and findings, and 7) Ethics and data handling. The items were formulated as questions for the guideline and presented in the first Delphi round for evaluation. An overview of the items included in the first Delphi round is shown in column 1 of Figure 2.

**Figure 2.**
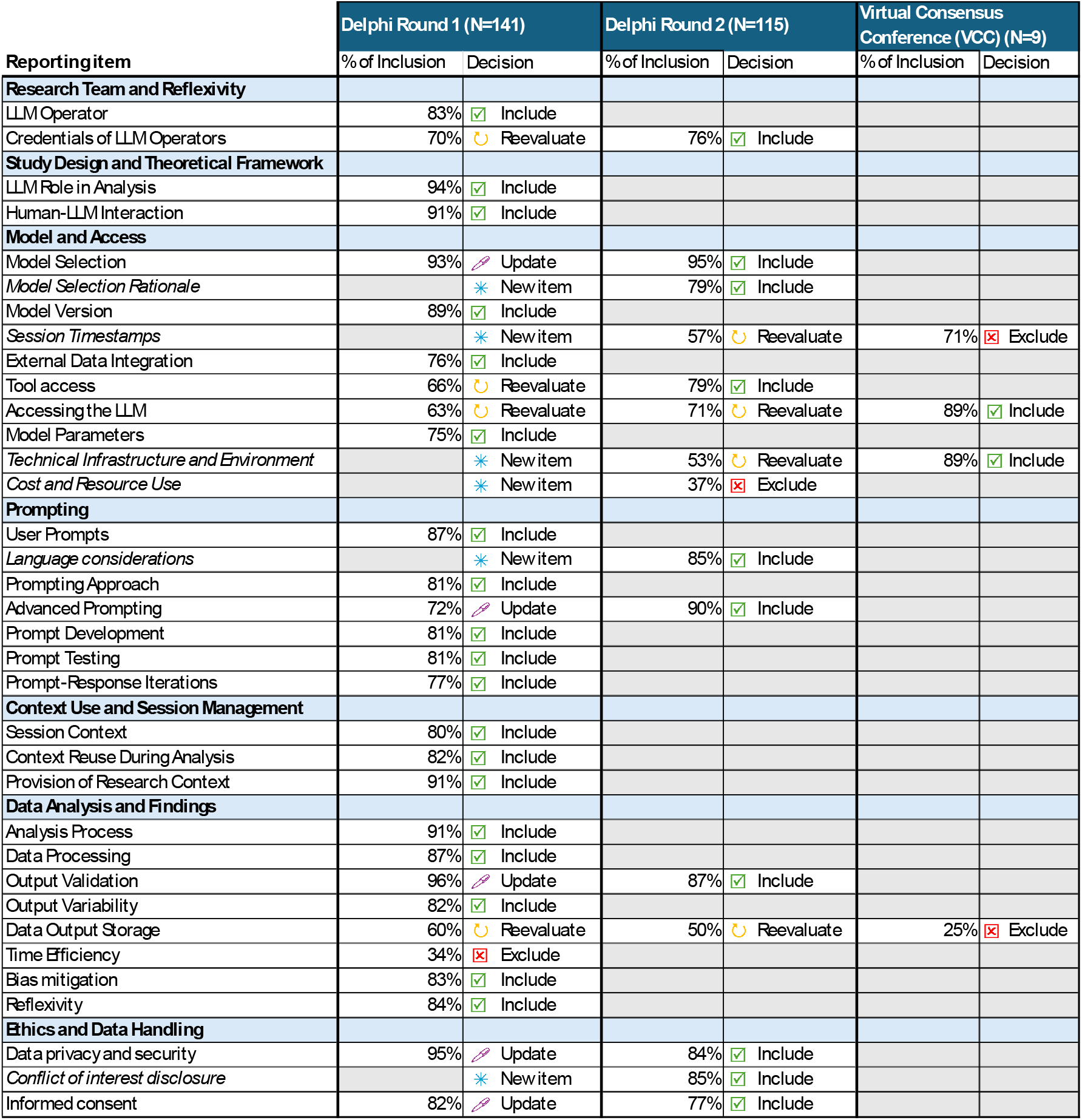
The first column lists all candidate items identified during the scoping review. Italicized elements indicate items added after Delphi Round 1 based on participant suggestions. Subsequent columns display voting results from the two Delphi rounds and the Virtual Consensus Conference (VCC). Participants rated each item on a 5-point Likert scale. Consensus for inclusion was defined a priori as >75% of respondents rating an item as 4 or 5 (agreement). Items with ≤50% agreement were excluded, while those with 51–75% agreement were revised and re-evaluated in the next round. In each round, participants could provide free-text feedback to suggest wording modifications or propose additional items.

### Delphi Consensus Process

We invited 1,455 experts from 67 countries; 145 participated in Round 1 resulting in an initial response rate of 9·7%. Participant demographics are shown in Figure 3.

**Figure 3.**
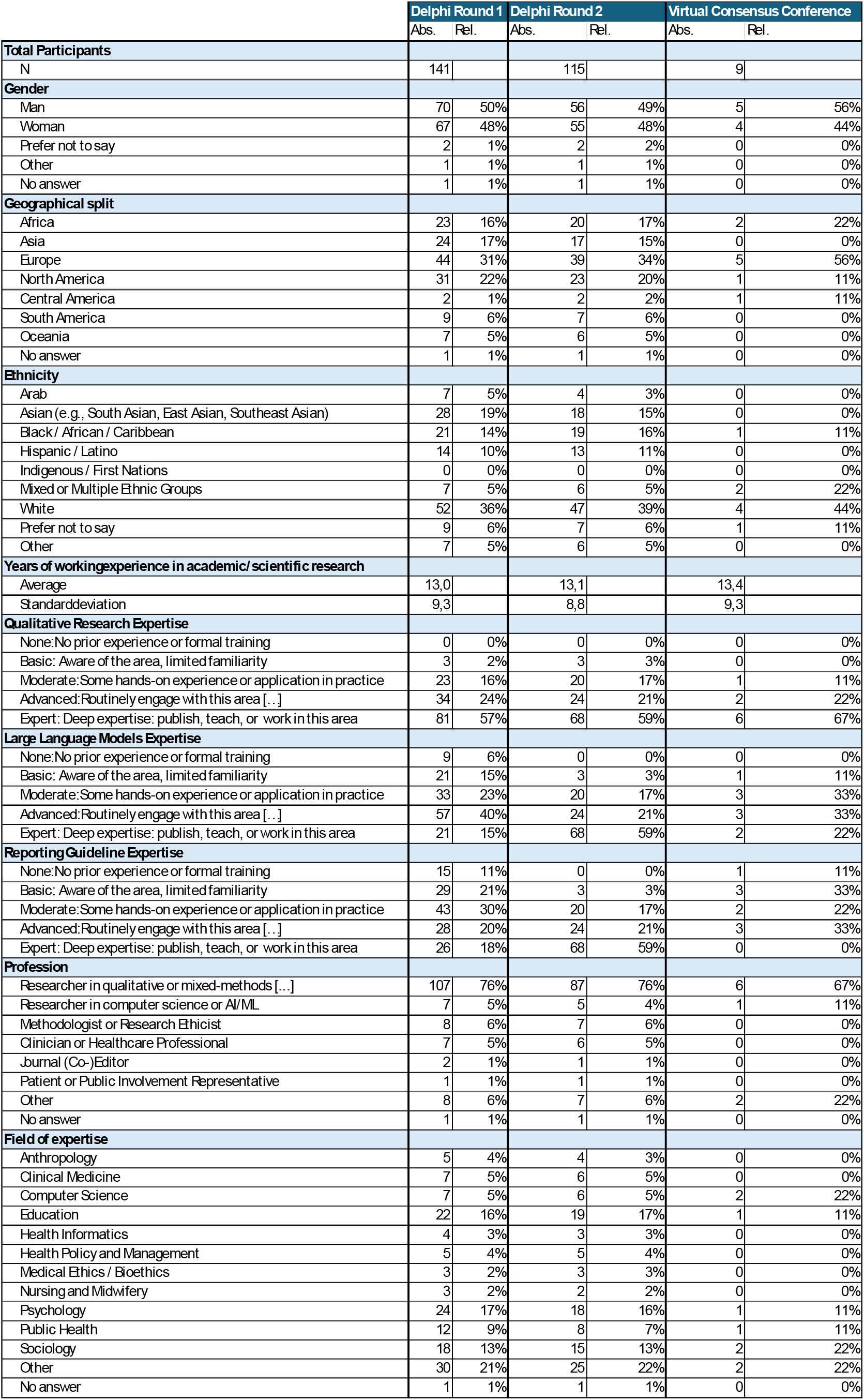
Participant demographics for the two Delphi rounds and the Virtual Consensus Conference are shown. The sample comprised experts from diverse countries, genders, and professional backgrounds.

During data cleaning, four response sets were excluded due to implausible answers (e.g., straight-lining) and unrealistically short completion times (The average time to answer the total survey was 19:36 min and the average time to rate the candidate items was 13:18 min). We excluded those that spend <2min on the rating of the candidate reporting items. In the second Delphi round, 115 responses were retrieved from the same participants, with no datasets requiring exclusion, resulting in an 81·6% follow-up response rate.

In Round 1, 19 of the initial items reached consensus (>75% agreement), four items required reevaluation in Round 2, one item was excluded, five items were suggested for alternative wording, and six new items were proposed in free-text comments. A complete list of free-text comments and how they were addressed in Round 2 is provided in Supplementary Material 2. The newly suggested items addressed session timestamps, conflicts of interest, reflections regarding the operating language used to communicate with the LLM, cost and resources, technical infrastructure, and model selection rationale. The latter was mentioned by several participants. Most wording revision suggestions aimed to clarify definitions or enhance comprehensiveness. For example, one participant proposed adding, “If multiple models were used, report each model and its role in the workflow,” to the item on model selection.

For the item on output validation, several participants recommended including a question regarding agreement between human coders and LLM outputs, and reporting statistical measures of inter-rater reliability (e.g., Cohen’s kappa or Fleiss’ kappa).

In Delphi Round 2 three of the six new items from Round 1 reached consensus. One item (cost and resource use) was rejected; participants noted that detailed reporting of hardware and software requirements and associated costs would provide limited additional value. All wording updates from Round 1 were approved in Round 2, and two items that had not reached consensus in Round 1 achieved the ≥75% agreement threshold.

As a result, four items remained undecided after Round 2 and were transferred to the Virtual Consensus Conference for final deliberation.

### Virtual Consensus Conference

All 115 participants from Delphi Round 2 were invited to the Virtual Consensus Conference to discuss the remaining open items. Nine experts attended. The panel agreed that the item on session timestamps when the LLM was used is not essential for the reporting guideline. Instead, reporting the LLM model version was deemed more relevant, which is part of a different item already. Session timestamps could only be included if authors are unable to identify the exact model version. There was broad consensus that the final checklist should include items detailing how the LLM was accessed and the technical infrastructure employed. Conversely, the item on data output storage was considered overly technical and not informative for the guideline.

The final checklist can be downloaded under Supplementary Material 3 as a an editable excel file. The excel document and further information will also be provided under https://coreq-llm.github.io.

### Scope of the Guideline

The primary focus of this guideline is reporting the use of LLMs in qualitative research, and the accompanying scoping review was similarly centered on LLMs. LLMs, e.g. Generative Pre-trained Transformer (GPT) family, are generative: they produce coherent, context-sensitive text and are increasingly used to assist qualitative workflows, including transcript summarization, exploratory theme generation and the production of synthetic responses that approximate participant viewpoints. These uses extend beyond simple classification.

By contrast, traditional natural-language processing (NLP)- and encoder-only models, e.g., bidirectional encoder representations from transformers (BERT)^29^, are primarily designed for contextual encoding and are commonly applied to tasks such as sentiment analysis, topic labelling and named-entity recognition. Although masked-token prediction can yield short spans of text, these models are not typically deployed to generate extended, interpretive prose.

During guideline development (Delphi preparation), it became apparent that the reporting principles also apply to other NLP models. We therefore asked panelists whether the guideline should extend beyond GPTs; agreement increased from 69% in Delphi Round 1 to 74% in Round 2. In the subsequent Virtual Consensus Conference, experts agreed that the principles are broadly applicable across NLP. Not all checklist items will be relevant in every case; authors may record items as ‘not applicable’ with a brief justification.

## Discussion

### General findings

The COREQ+LLM initiative represents the first effort to adapt qualitative research reporting standards to an era in which human interpretation increasingly coexists with algorithmic reasoning. LLMs are not neutral tools, they influence what can be known, how meaning is constructed, and whose voice is represented in qualitative analysis. By making their use visible and reportable, COREQ+LLM contributes to preserving interpretive rigor and ethical accountability in a rapidly changing research landscape. Across three methodologically rigorous phases, we identified, refined, and validated a set of reporting items that capture essential methodological, ethical, and epistemological considerations associated with LLM use in qualitative research.

The scoping review documented substantial gaps in existing reporting and guided item generation. Through a two-round Delphi process and a Consensus Conference international experts from qualitative research, AI informatics and publishing consensus was reached for 31 items organized into 7 categories.

By offering standardized guidance, COREQ+LLM aims to improve transparency, reproducibility, and credibility of qualitative research involving LLMs. The checklist enables reviewers and editors to assess whether LLM-based tools were applied responsibly, promotes explicit consideration of ethical and epistemic risk, and provides a shared vocabulary for describing human–LLM collaboration. Beyond its tility for manuscript reporting, the checklist may influence research design and reviewer training, promoting reflective and responsible innovation in digital qualitative methods.

### Interpretation of consensus dynamics

While the Delphi process achieved high overall agreement, variation across items reflected differing epistemic orientations among panelists. Experts rooted in traditional qualitative methodology often emphasized reflexivity, human interpretive control, and contextual transparency, whereas participants with more technical backgrounds prioritized model specification, prompt documentation, and reproducibility. These contrasting emphases underscore that LLM use in qualitative research bridges two methodological cultures: interpretive and computational. The resulting COREQ+LLM items therefore balance interpretive rigor with technical precision, promoting both interpretive depth and procedural transparency. This highlights that reporting LLM use is not solely about documenting software parameters but about articulating the evolving relationship between human researchers and LLMs.

### Distinction from other guidelines

COREQ+LLM extends the original COREQ checklist. Whereas COREQ covers the use of any software as a single reporting item, COREQ+LLM specifies how, when, and why LLMs are used and how their probabilistic nature affects interpretive validity. Studies using an LLM-assisted qualitative approach should report against original and extension checklist, where applicable. Existing AI-oriented reporting frameworks, such as CONSORT-AI, SPIRIT-AI, and TRIPOD-LLM, focus primarily on quantitative use cases. COREQ+LLM therefore fills a critical methodological gap, providing qualitative researchers with a contextually grounded yet flexible reporting structure aligned with interpretive traditions. Although we focused on generative LLMs, the consulted experts in this study agreed that the COREQ+LLM guideline is applicable to other NLP-techniques and LLM-architectures as well, including models such as BERT.

### Limitations and future research

Several limitations should be acknowledged. First, the scoping review was restricted to English-language publications and may therefore not capture regional or non-English methodological innovations. Second, although the Delphi panel was international and multidisciplinary, it was skewed towards researchers from Europe and North America with backgrounds in qualitative research; perspectives from the Global South, where infrastructural and linguistic barriers to LLM use differ, and technical viewpoints from AI specialists may have been underrepresented. The panel for the Virtual Consensus Conference was rather small with only 9 participants which may have resulted in some selection bias. However, the limited sample enabled each participant to voice their perspective, resulting in a rich and engaging discussion.

Finally, while the Delphi method supports structured expert consensus, such consensus reflects collective judgment rather than empirical validation; future studies should evaluate the guideline in a real world setting and examine whether adherence to COREQ+LLM enhances reporting quality and methodological rigor in practice.

To facilitate correct implementation of the reporting guideline, the authors plan to develop and disseminate supplementary background materials and practical guidance on its use.

In line with emerging expectations for transparency in AI-assisted research across disciplines, COREQ+LLM contributes to a broader cultural shift toward traceable and auditable human–AI collaboration in knowledge production. Its value lies not only in improving how studies are reported but in shaping how they are conceived and conducted.

## Conclusions

The development of COREQ+LLM marks an important step toward responsible and transparent integration of LLMs into qualitative health research. Through systematic evidence synthesis and expert consensus this guideline provides a practical tool for ensuring methodological rigor in an era where human interpretation increasingly intersects with machine-generated insight.

## Data Availability

All data produced in the present study are available upon reasonable request to the authors

## Acknowledgments

This study is supported by the internal research fund of the Faculty of Health, School of Medicine, Witten/Herdecke University, Germany (Project IFF 2024-80). Julian Frings receives financial support through a doctoral scholarship from the German Academic Scholarship Foundation. The funders will play no role in study design, data collection, analysis and interpretation of data, or the writing of this manuscript.

The authors want to thank all experts who participated in the consensus process of this project, namely:

PhD Abderrahmane Benlahcene - College of Commerce and Business Administration, Dhofar University; MBA Abisha Kampira - Tshwane University of Technology; Dr. Abraham Mhaidli - Max Planck Institute for Security and Privacy; MSc. Adekunle Ajibewa - Obafemi Awolowo University, Ile Ife, Nigeria; PhD (Education) Ahmad Zulfadhli Khairuddin - Universiti Teknologi MARA; MS Alex Liu - University of Washington; Lecturer at the Faculty of Arts, Helwan University, Faculty of Arts, Department of Sociology Ali Aboalhassan - Helwan University, Faculty of Arts, Department of Sociology; MSc Aminu Yakubu - Center for Bioethics and Research, Ibadan, Oyo State, Nigeria; MAP (Master of Applied Psychology) Amy S.

G. Lee - University of Melbourne; Associate professor, MD, MSc, MPhil, MS, FACS Anaïs Rameau - Department of Otolaryngology - Head & Neck Surgery, Weill Cornell Medicine, New York, NY, USA; Prof. André Fringer - Zurich University of Applied Sciences ZHAW; MPhil Andrew Chimatira - Research Methods International; PhD Angélique Herrler - Charité - Universitätsmedizin Berlin, Institut für Medizinische Soziologie und Rehabilitationswissenschaft; PhD Antoni Casasempere Satorres - CualSoft Consulting; Prof. Antonio M. Nogués - Universitas Miguel Hernández; PhD Ashish Vazirani - Shiv Nadar University; Ph.D. Assel Csonka-Stambekova - n/a; DPhil Baptiste Vasey - University of Oxford; Dr/PhD Bethany Morgan Brett - City St Georges, University of London; Dr. Bjorn de Koning - Erasmus University Rotterdam; Associate Professor Bornface Chinene - University of Rwanda, College of Medicine and Health Sciences, Department of Medical Imaging Sciences, Remera Campus; Prof. Bruno Falissard - Université Paris-Saclay; PhD Burcu Şentürk - Assoc.Prof.; PhD Byron Qually - research + DESIGN (Pty) Ltd.; MA Caitlin Tefft - University of Wisconsin - Madison; PhD Caterina Fugazzola - The University of Chicago; MD, MS Charles Kahn - University of Pennsylvania; MSc Chidi Hope Agbaraji – Littleseed Concepts and Label Limited; Dr. Corina Güthlin - Institute of General Practice /Goethe-Universität Frankfurt; PhD Daniela Castellanos-Reyes - North Carolina State University; PhD Daniele Proverbio - University of Trento; A/prof Danielle Hitch - Deakin University; Prof. Dr. Débora Cecilia Chaves de Oliveira - Universidade do Estado do Rio de Janeiro (UERJ); Doctoral Scholar (Pursuing PhD) Denix Sebastian Indian Institute of Management Tiruchirappalli; PhD Dilara Özel - Middle East Technical University; MSc Eda Şen Bircan - -; MSc, MA, BSc Edwin Okoyo - TISL; PhD. Eli Malvaceda - Universidad San Ignacio de Loyola; PhD Elias Balt - Research department, 113 Suicide Prevention; Amsterdam Public Health school, Amsterdam University Medical Center; Assoc. Dr. Elif Kus Saillard - Founder; PhD Elise van der Mark - Senior researcher at IISH, Amsterdam the Netherlands; MD MPH Eric Poon - Duke University School of Medicine; Phd. Ciencias políticas y Sociales, Orientación en Ciencia Política Erika Tapia Nava - Instituto de Investigaciones Dr. José María Luis Mora/ Facultad de Ciencias Políticas y Sociales, UNAM; MSc Erkan Eyrikaya - Department of Clinical Psychology, Graduate School of Social Science, Ankara University, Ankara, Türkiye; PhD Esengul Elibol - Istanbul Bilgi University; MSc, EMBA Eugene Swinnerstone Miheso - Ernest Cook University; MSc (Bioethics) Evelaynne Hort - ATLAS.ti Professional Junior Trainer; Prof Farah Magrabi - Australian Institute of Health Innovation; PhD Filip Schmidt - Adam Mickiewicz University Poznan; MSc. Francisco Freitas - Universidade de Coimbra; PhD Gonzalo Seid - CONICET (Argentina) & Instituto de Investigaciones Gino Germani de la Universidad de Buenos Aires; PhD Hakan Tarhan - TOBB University of Economics and Technology; PhD Hannah Frith - University of Surrey, UK; M.A.Ed. Hilmi Tahrir - Universiti Tun Hussein Onn Malaysia, UTHM; Prof Hongfang Liu - University of Texas Health Science Center at Houston; PhD Idowu Omisile - Obafemi Awolowo University, Ile-Ife, Osun State; PD Dr. Irina Jarvers - Department of Child and Adolescent Psychiatry and Psychotherapy, University of Regensburg; Prof. / Ph.D. Jacquie Kurland - University of Massachussetts; PhD Janet - Global Health Mentorships/ University of Bern; MSc Jason Robert Rameshwar; Professor Jegede Ayodele Samuel - University of Ibadan; MSc Jemal Seid Ahmed - Qualitative research Analyst and Junior Atlas.ti trainer; Prof Jiang Bian - Indiana University; Prof/PhD Jiang Bian - Indiana University; Dr., M.Pd Joko Soebagyo - Universitas Muhammadiyah Prof. Dr. Hamka; PhD José Laurián Ramírez Díaz - Instituto Tecnológico de Costa Rica; PhD Jose M Valderas - National University of Singapore; PhD Julia Trzcińska - University of Wrocław; prof Karel GM Moons - Julius Center for Health Science and Primary Care, UMC Utrecht, Utrecht University, Utrecht, The Netherlands; PhD Kathryn Wheeler - University of Essex; PhD, RN Kelli DePriest - RTI International; MSc Kousik Mallick - PhD Scholar; MSc Kübra Annac - Universität Witten/Herdecke; PhF Ladislav Toušek - University of West Bohemia; PhD Lief Esbenshade - University of Washington; PhD Lois Player - University of Bath; PhD Lynn Monrouxe - The University of Sydney; PhD Marya Bekova - Nazarbayev University; M.S., M.P.H Maximo Prescott - University of California, San Diego; Research Associate Professor & Principal Scientist Mayank Kejriwal - University of Southern California; Prof/PhD Melanie Calvert - University of Birmingham; MSc Michelle Stage - University of Rhode Island; PhD Milan Stančić - University of Belgrade - Faculty of Philosophy; Dr Mohammad Alsubaie - Ethar of knowledge for Surveys and Studies; PhD Mostafa Amini-Rarani - Social Determinants of Health Research Center, Isfahan University of Medical Sciences, Isfahan, Iran.; PhD Candidate Nicole Reinsdorf - Universität Potsdam; PhD Nove E. Variant Anna - Universitas Airlangga; PhD Olivia Brown - University of Bath; PhD Oluwafunke Aduwo - Department of Sociology, University of Ilesa, Ilesa, Osun State.; MSc Orofin - Abigail; PhD Oscar Castro - Universidad Distrital Francisco José de Caldas; Dr. Oshri Bar-Gil - Bar-Ilan University; PhD Pamela Castillo Mardones - Facultad Ciencias Sociales, Universidad de Concepción, Chile; PhD Piotr Binder - Institute of Philosophy and Sociology, Polish Academy of Sciences; PhD Candidate Qiming Xie - Nanjing University of Science and Technology; Professor, M.D., Ph.D. Rene Weber - University of California Santa Barbara; Dr/PhD Richard Douglas Kamara - University of Johannesburg; PhD Rimke Bijker - Department of Psychology and Neuroscience, Auckland University of Technology, Auckland, New Zealand; Ph.D. Ruth Slotnick - Bridgewater State University; Ph.D Salime Goharinezhad - School of Health and Care Management, Arden University, Arden House, Middlemarch Park, Coventry CV3 4FJ, United Kingdom.; PhD Salome Kasimba - Research Consultant; Assistant Professor of Neurology/ PhD Sandhya Seshadri - University of Rochester; PhD Sandra Steyn - Research4You; Da Vinci Institute; MS Clinical Psychology Sara Haqqi - Muhammad Ali Jinnah University; PhD Sarah Yeo - University of Arizona; PhD Sidikat Shitu - Ibrahim Badamasi Babangida University, Lapai, Niger State, Nigeria.; PhD Simone Rodda - Auckland University of Technology; PhD Soleh Hasan Wahid - Universitas Islam Negeri Kiai Ageng Muhammad Besari Ponorogo; Dr. Stefanie Panke - University of North Carolina at Chapel Hill; PhD Steven Wright - University of Edinburgh; Dr. Susanne Sackl-Sharif - University of Graz; MD, PhD, MHPE Takeshi Kondo - Nagoya University Hospital; MSc, EMPH Till Neugebauer - Witten/Herdecke University, Faculty of Health, School of Medicine, Health Services Research; PhD. Titilope Abosede Oderinwale - International Centre for Educational Evaluation, Institute of Education, University of Ibadan, Nigeria; Dr. Tuğçe Yerlitaş - ISM University of Management and Economics; PhD Education Ulises Alejandro Duarte Velazquez - Escuela Normal Oficial de Irapuato; MD Uriel Fennig - 1 - Department of Neurology, Sheba Medical Center at Tel Hashomer, Ramat Gan, Israel. 2 - Faculty of Medical and Health Sciences, Tel Aviv University, Tel Aviv, Israel; MSc Urs Fichtner - Section of Health Care Research and Rehabilitation Research, Institute of Medical Biometry and Statistics, Medical Faculty and Medical Center - University of Freiburg; MSc Victor Iyiola - University of Medical Sciences, Ondo; MA Vilma Sukackė - Kaunas University of Technology; MD Walter Mathis - Yale School of Medicine; MSc William Sebunje - Mildmay research and Ethics Committe; Associate Prof., MD, PhD, FRCPC, FCPA, FAPA Yanbo Zhang - University of Alberta; Ph.D. Yang Yang - Texas Christian University; B.S. Yongjie Sun - The School of Psychology and Cognitive Science, East China Normal University; Department of Social Development, New York University Shanghai; Professor Yufei Yuan - McMaster University; PhD Yvonne Leung - Northeastern University- Toronto; DBA, Doctorate of Business Administration Zahraa Berro - Independent researcher in qualitative methods; Prof. Dr. Zhaohui Su - Southeast University.

## Availability of data and materials

The study protocol is available here: doi: 10.2196/78682. The de-identified data sheet for each Delphi Round used to generate the results in this paper can be provided by the corresponding author upon reasonable request.

### Declaration of generative AI and AI-assisted technologies in the manuscript preparation process

During the preparation of this work the authors used OpenAI’s ChatGPT (GPT-5o) in order to improve grammar, spelling, and punctuation of this manuscript. After using this tool/service, the authors reviewed and edited the content as needed and take full responsibility for the content of the published article.

## Author Contributions

Leonard Fehring: Conceptualization, Methodology, Writing – Original draft and Review & Editing, Data analysis, Supervision, Project Administration.

Julian Frings: Methodology, Data collection and analysis (Delphi survey), Data Curation.

Paul Rust: Methodology, Writing - Review & Editing, Data Curation.

Christian Kempny: Writing - Review & Editing, Data analysis (Literature Review) Petra Thürmann: Methodology, Supervision, Writing - Review & Editing, Validation. Emre Tamerler: Data collection and analysis (Delphi survey and Virtual Consensus Conference), Data Curation.

Sven Meister: Methodology, Writing - Review & Editing, Validation.

