## Supplementary Material 1 for "Reporting of Qualitative Research Using Large Language Models (COREQ+LLM): A Delphi-based Extension of the COREQ Reporting Guideline"

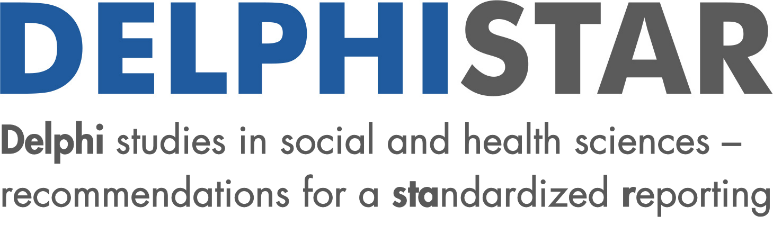

**Delphi studies in social and health sciences – recommendations for an interdisciplinary standardized reporting (DELPHISTAR).**

| **Topic** | **Section** | **Item** | **Checklist Item** | **Location where item is reported** | **Exemplary wording** |
| --- | --- | --- | --- | --- | --- |
| **I**  **Title and Abstract** |  | 1 | Identification as a Delphi study in the title | Yes (pg.1) | What is a public health intervention? Results of a Delphi study. |
|  |  | 2 | Identification as a Delphi study in the abstract | Yes (pg.1) | A Delphi study was selected to answer the research question. |
|  |  | 3 | Structured abstract | Yes (pg.1) | e.g., background, method, results and discussion |
| **II**  **Context** | **Formal** | 4 | Information about the sources of funding | Yes (pg.13) | The Delphi study was funded by [SOURCE]. |
|  |  | 5 | Information about the team of authors and/or researchers (e.g., discipline, institution) | Yes (pg.6) | The Delphi study was conducted by an interdisciplinary team with representatives from medicine, public health, and health promotion. |
|  |  | 6 | Information about method consulting | Yes (pg.6) | The study group was advised by experts from [INSTITUTION] regarding statistics.  Or:  No consulting in regard to method took place. |
|  |  | 7 | Information about the project background | Yes (pg. 4f) | The Delphi study was part of a mixed-methods study on [AIM]. |
|  |  | 8 | Information about the study protocol | Yes (pg. 4) | The study protocol is available at [LINK]. |
|  | **Content** | 9 | Justification of the chosen method (Delphi) to answer the research question | Yes (p.4) | The Delphi method is suitable for answering the research question because it systematically gathers the judgments of different expert groups and can identity agreement and disagreement. |
|  |  | 10 | Aim of the Delphi study (e.g., consensus, forecasting) | Yes | The aim of the Delphi study is to find consensus on criteria to define a public health intervention. |
| **III**  **Method** | **Body & Integration of knowledge** | 11 | Identification and elucidation of relevant expertise, spheres of experience, and perspectives (e.g., theory, practice, affected groups, disciplines) | Yes (pg.6) | The experts represent the sciences and clinical practice because [REASON]. |
|  |  | 12 | Handling of knowledge, expertise and perspectives which are missing or have been deliberately not integrated | Yes (pg.6) | If it is not possible to recruit experts specialized in [AREA], this will be openly communicated to the other experts during the Delphi study. |
|  |  | 13 | Basic definition of expert^1^ | Yes (pg.6) | A person who has been active in the area for at least [NUMBER] years is considered to be an expert. |
|  | **Delphi variant and modifications** | 14 | Identification of the type of Delphi variant and potential modifications (e.g., classic Delphi, real-time Delphi, group Delphi) | Yes (pg.6) | A classic Delphi study was used [LITERATURE REFERENCE]. |
|  |  | 15 | Justification of the Delphi variant and modifications, including during the Delphi study, if applicable | N/A | If the willingness to participate clearly decreases between the first and second round, a third round will not be held. |
|  | **Sample of experts** | 16 | Selection criteria for the experts (per round, per expert group if applicable) | Yes (pg.6) | All of the experts who met the definition were invited to the first round.  All of the experts who completed the previous round were invited to participate in the subsequent round. |
|  |  | 17 | Identification of the experts | Yes (pg.5-6) | The experts were identified based on publications in [DATABASE]. |
|  |  | 18 | Information about recruiting and any subsequent recruiting of experts | Yes (pg.6) | The experts were informed about the Delphi study and invited to participate. |
|  | **Survey** | 19 | Elucidation of the content development for the questionnaire^2^ | Yes (pg.5) | The questionnaire was developed based on the results of systematic reviews [LITERATURE REFERENCE]. |
|  |  | 20 | Description of the questionnaire (content and structure) | Yes (pg.6) | The questionnaire was divided into three segments on [TOPICS]. The statements made in the questionnaire were evaluated using standardized items, with the option to comment in free-text boxes. |
|  | **Delphi rounds** | 21 | Number of Delphi rounds | Yes (pg.6) – impliziert weil wir jede Runde beschreiben | Three Delphi rounds were held. |
|  |  | 22 | Information about the aims of the individual Delphi rounds | Yes (pg.6) | The first Delphi round focused on exploring relevant aspects. These aspects were then presented to the experts in the second Delphi round for standardized evaluation. |
|  |  | 23 | Disclosure and justification of the criterion for discontinuation | Yes (pg.6) | The number of rounds was defined in advance to be a maximum of three rounds. |
|  | **Feedback** | 24 | Information about what data was reported back per round | Yes (pg.6) | In terms of feedback, we shared the statistical results plus the summary of the open responses. |
|  |  | 25 | Information on how the results of the previous Delphi round were fed back to the experts surveyed (e.g., via frequencies, mean values, measures of dispersion, listing of comments) | Yes (pg.6) | Mean values, standard deviations and percentage frequency distributions were reported. |
|  |  | 26 | Information on whether feedback was differentiated by specific groups (e.g., by field of expertise, institutional affiliation) | Yes (pg.6) | The feedback was aggregated across all expert groups. |
|  |  | 27 | Information about how dissent and unclear results were handled | Yes (pg.6) | The results showing dissent were presented again for evaluation in the next Delphi round. |
|  | **Data analysis** | 28 | Disclosure of the quantitative and qualitative analytical strategy | Yes (pg.6) | The quantitative items were descriptively analyzed. The open-ended items were analyzed using thematic analysis [LITERATURE REFERENCE]. |
|  |  | 29 | Definition and measurement of consensus | Yes (pg. 6) | Consensus was defined as percentage agreement, meaning that agreement was assumed if at least 80% of the respondents agreed on an item. |
|  |  | 30 | Information on group-specific analysis or weighting of experts (e.g., theory vs. practice, discipline-specific analysis) | N/A | In the analysis, the mean values for percent agreement are weighted for each expert group in terms of the number of group members. |
| **IV**  **Results** | **Delphi process** | 31 | Illustration of the Delphi study (e.g., in a flow chart) | Yes (pg. 8 und 10) | A summary of the Delphi study is illustrated in a flow chart (Figure 1). |
|  |  | 32 | Information about special aspects during the Delphi study (e.g., deviations from the intended approach with justification) | N/A | During the Delphi study the political discussion mentioned climate change and the effects on health. It is possible that this influenced the experts' responses. |
|  |  | 33 | Number of experts per round (both invited and participating) | Yes (pg. 7) | The number of experts participating in the first Delphi round was [NUMBER], and the number of experts in the second round was [NUMBER]. This corresponds to a response rate of [NUMBER]% in the first round and [NUMBER]% in the second round. |
|  | **Results** | 34 | Presentation of the results for each Delphi round and the final results | Yes (pg. 8-10) | In the first Delphi round [NUMBER]% of the experts agreed, in the second [NUMBER]%, and in the third [NUMBER]%. |
| **V Discussion** | **Quality of findings** | 35 | Highlighting the findings from the Delphi study | Yes (pg. 11-12 beim Beginn der Diskussion) | The central findings can be summarized as follows: [STATE FINDINGS]. |
|  |  | 36 | Validity of the results (e.g., transferability of the findings) | Yes (pg.12) | The results are not transferable to other countries due to different legal regulations. |
|  |  | 37 | Reliability of the results (e.g., split half, inter-rater reliability) | Yes (pg. 12) | The responses in the free-text comments were analyzed by two independent reviewers [SPECIFY]. |
|  |  | 38 | Reflection on potential limitations (e.g., number of experts, response bias) | Yes (pg. 12) | The results are to be viewed critically with regard to the composition of the panel because [REASONS]. |
