## Supplementary Material 2 for "Reporting of Qualitative Research Using Large Language Models (COREQ+LLM): A Delphi-based Extension of the COREQ Reporting Guideline"

| Comments: Do you have suggestions for additional reporting items or revisions to the items listed above? | Response / How it is implemented |
|---|---|
| Not at this time, the list will seem exhaustive. | Thank you for your feedback. |
| The issue of reflexivity is key and also the importance of comparing themes obtained after analysing data without AI assistance before and what emerges after AI assistance. It would be great to identify how AI is shaping our thinking as the researcher is considered an instrument in qualitative research .<br>My concern is how to make reporting this coincise and not cumbersome | Thank you for highlighting the importance of reflexivity and comparative analysis between AI-assisted and traditional qualitative methods.<br><br>These critical aspects are addressed in our existing reporting items "Reflexivity" and "Human-LLM Interaction". We share your concern about maintaining conciseness in reporting and have designed the guidelines to balance thoroughness with practical implementation. |
| Additional Suggestions for Reporting Items<br>I recommend adding the following:<br>1. Version Control & Timestamps. Include timestamps for when prompts were run and which model version was active at that time (especially for models with frequent updates).<br>2. Cost and Resource Use. Report approximate computational cost (e.g., API costs, compute time) to inform feasibility for other researchers.<br>3. Failure Modes and Limitations. Describe instances where the LLM failed to produce useful output or required correction. This adds depth to methodological reflection.<br>4. Human Coding Comparison. If manual coding was done in parallel, report level of agreement (e.g., kappa, percentage) between human and LLM outputs.<br>5. Data Retention Policies. Clarify whether the LLM provider retains input data and what steps were taken to prevent storage (e.g., disabling chat history, using enterprise APIs with data protection). | We greatly appreciate your detailed suggestions for additional reporting items.<br><br>Based on your recommendations, we have incorporated the following new items: "Session Timestamps" and "Cost and Resource Use".<br><br>We have expanded the existing items "Output Validation" and "Data privacy and security" section to include failure analysis and limitations, interrater-agreement, and privacy concerns. |
| I think some of the items are largely related to one another. I.e., multi-turn interactions will overlay with whether or not the interaction took place in a clean, new environment or if there were measures already included in the thread. I think this is a really excellent idea to be developing this - very important work, thank you. | We acknowledge that certain aspects, such as multi-turn interactions and conversation environment management, are indeed interconnected. We appreciate your support for this work. |
| I think it should ask how many and which Llama are used or if multiple LLMs are used. Also, I think it would be a great idea to ask if the AI was given any coding dictionary or scheme for coding. To check for inter-coder agreement I recently uploaded a scheme for coding and a coding dictionary and asked ChatGPT to code the same interview so that I can see if AI was assigning same codes and themes as human coders. | Thank you for your valuable suggestions regarding multiple model usage and coding scheme integration.<br><br>We have revised the "Model Selection" item to address scenarios involving multiple LLMs, adding the requirement: "If multiple models were used, report each model and its role in the workflow."<br><br>The integration of coding dictionaries and schemes is addressed through the "Advanced Prompting" and "LLM Role in Analysis" items, we have revised to include the documentation of materials provided to the LLM including codebooks.<br><br>Additionally, we have added "Interrater-agreement" under "Output Validation" to capture the reliability assessments you described. |
| I would add WHY a certain model was chosen upon others (various models are known to present different stability, bias, geopolitical, or cost properties. Specifying the reason of the model selection would enhance reproducibility and possibly interpretability) | Thank you. Following your recommendation, we have added a new reporting item "Model Selection Rationale" that requires researchers to document their reasoning for choosing specific models. |
| I think it is important that artificial intelligence is used as a peer in qualitative research. Researchers remain the primary instrument for data collection and analysis. Even if AI can produce better results, it will never be the main instrument in qualitative research. In addition to models such as Grok or ChatGPT, analysis options such as those offered by MAXQDA or Tailwind should also be considered for COREQ. It should also be noted whether only one tool or several in combination were used as an analysis path based on their strengths, for example ChatGPT => NotebookLM => MAXQDA => Tailwind. | We have revised the "Model Selection" item to accommodate multiple tools and workflow combinations, including the requirement to report each tool's specific role when multiple systems are employed. |
| LLM's speed and costs are highly dependent on the hardware and LLM providers. Many LLMs can be hosted locally to enhance privacy and reduce cost. But it will take a lot of prompting and examples. Some experts can fine-tune their open source LLMs to conduct the qualitative data analyses. | We have incorporated a new item "Cost and Resource Use" and expanded the existing "Data privacy and security" item to address costs as well as as privacy considerations. |
| No | Thank you for your feedback. |
| I believe AI agents will become widely used in the coming years. In that case, I think it will also be necessary to state whether the source code of the entire system is disclosed. | We have revised the "Model Selection" item to include whether the model was an open- or closed-source model. |
| Consider including elements that address transparency regarding the limitations of the model, reporting on associated costs and resources, and evaluating whether researchers have received training in AI ethics. Reflecting on how the use of large language models (LLMs) influenced findings, as opposed to traditional qualitative methods, may offer valuable insights. Additionally, when applicable, it might be insightful to consider participants' perspectives regarding LLM involvement. Encouraging the inclusion of reproducibility materials such as scripts, prompts, and outputs should also be a priority, when ethical considerations allow. | Thank you for your comprehensive suggestions.<br><br>We have addressed your recommendations through several items: "Cost and Resource Use" for transparency regarding associated costs; operator training in AI ethics is covered under "LLM Operator" and "Credentials of LLM Operators"; model limitations and comparison with traditional methodsare addressed in the "Reflexivity" item.<br><br>Your emphasis on reproducibility materials aligns with our requirements for sharing scripts and prompts in supplementary materials where ethically appropriate. |
| None | Thank you for your feedback. |
| I think it would be important to also report on what was not delegated to the LLM. For example, what did the researchers do without the use of the LLM? Did they leave the interpretation to human researchers, or did the human researchers only validate interpretations? The authorship boundaries should also be addressed. It would be important to know in which phases of the analysis process the LLM was used and how it was used. | We appreciate your observation about delineating human versus LLM responsibilities in the research process. The items "LLM Role in Analysis," "Human-LLM Interaction," and "Output Validation" are designed to capture the specific tasks assigned to the LLM, with the understanding that non-specified tasks remain under human researcher control. This approach provides clarity about task delegation while maintaining transparency about the boundaries of AI assistance in the analytical process. |
| Fields such as data sciences are becoming fundamental sciences—and indeed, we can already say that this is happening. Therefore, having someone from data science on the research team does not necessarily add positive value to the study in terms of reliability or in any other respect. However, the opposite might be true: for research teams without a formal diploma in this area, this could potentially create problems. On the other hand, there are countless examples showing that good programmers often do not hold formal degrees. :D<br><br>In addition, it may be important to include some questions in appropriate places that address more technical aspects such as cross-validation or external validation.<br><br>Regarding bias, some items could also cover the detailed description of the sample. There are numerous studies in the field where the age and gender distribution of the dataset is not reported. In this regard, I believe additional questions should be included.<br><br>If programming was used, the specific programming language, libraries, and exact version numbers, along with access dates, should also be reported. This would ensure transparency, as errors can sometimes occur between different versions. | Thank you for your observations about credentialing and technical specifications.<br><br>We have added "Interrater-agreement" to address validation concerns and "Technical Infrastructure and Environment" to capture the technical details you mentioned.<br><br>In case where a model was fine-tuned with a specific dataset was used, we expect this to be reported under the item "Model selection".<br><br>The bias and misinformation concerns you raised are addressed through our "Bias Mitigation," "Output Validation," and "Reflexivity" items. |
| You may also include criteria related to qualitative coding validation, where the LLM can be treated as one "coder." This enables the operator to calculate and report on:<br>Cohen's Kappa – for two raters (e.g., LLM vs. human coder).<br>Fleiss' Kappa – for more than two raters (e.g., LLM plus multiple human coders). | We appreciate your specific recommendation regarding validation metrics. We have incorporated "Interrater-ragreement" as an extension of the "Output Validation" item that explicitly requires documentation of inter-rater reliability measures, including Cohen's Kappa for two raters and Fleiss' Kappa for multiple raters, when treating the LLM as a coder in the analytical process. |
| No I don't | Thank you for your feedback. |
| Perhaps the type of qualitative analysis and associated epistomology etc and whether the LLM was advised of the approach. Also whether there was a specific prompt for data anlaysis approach (e.g., TA, QCA, GT). maybe include in triangulation running the models with different LLMs. | Thank you for your suggestions regarding epistemological frameworks and methodological approaches. The underlying qualitative methodology is addressed in the original COREQ guidelines, while the communication of these approaches to the LLM is captured under "User Prompts." The triangulation with different LLMs you mentioned is addressed in our "Prompt Testing" item. |
| This is timely research. Let me know how I can be part of this ongoing investigation.<br><br>My suggestion and question ...<br><br>We need to address codebook development and refinement in context to the particulate research. Another aspect is auditable traceability- fron a database standpoint- we need to know how wual raw was processed to where it is.Lastly, how can member checking be effected in the LLM for qual reporting be effected so that the LLM reporting is not to the researcher but the participant and what will be the ethical steps for doing so? | We have expanded the existing item "Data privacy and security" as a new item to address the auditable traceability concerns you raised. The codebook development and refinement processes are covered under existing items for prompting and analysis documentation. Your question about member checking with LLM-generated outputs raises significant ethical considerations that warrant further research and development beyond the current scope of these reporting guidelines. |
| Would there be an ethical issue if participants were told the research would use LLM? Is it likely that they may object? (Just asking) | Thank you for raising this consideration.This is covered by the exisiting item "Informed consent". |

| | |
|---|---|
| n/a | Thank you for your feedback. |
| I believe inclusion of the particular type of LLM should not be very explicit. | Thank you for your feedback. We have considered your perspective on the specificity of LLM disclosure and believe that transparency regarding the specific models used remains essential for research reproducibility and evaluation. The decision to keep this aspect in the final guideline will depend on the outcome of the Delphi process. |
| Yes. I suggest adding the following item: In cases of discrepancies between human coders and LLMs, what decision rule or rationale was applied to resolve them? | We appreciate your suggestion regarding discrepancy resolution. This has been incorporated into the new "Interrater-agreement" aspect under "Output Validation," which explicitly requires documentation of decision rules and rationales applied when resolving differences between human coders and LLM outputs. |
| The nature and role of the different operators may be more clearly defined. Operating the LLM can in my view be as narrow as providing the model with the prompts and data that have been decided upon in advance by the research team. A research team could even develop an intuitive front end for their LLM, making operating the model a simple administrative task. I feel that it is more important how the research team came to select their model, developed and tested their prompts, validated their data, etc. | We have added "Model Selection Rationale" to document the decision-making process behind model selection. Our items "LLM Operator" and "Credentials of LLM Operators" are designed to capture the full spectrum of roles and expertise involved in LLM implementation. |
| I think a lot of these questions assume that the human is in control and know how the models are working. But particularly if you are using a custom built tool (like in MAXQDA and NVivo), you are not aware of what's happening but that may be ok because there may be some trust that the prompt designers have developed this tool with qualitative research in mind. But with the big LLMs like Chat GPT, I have no idea how it works and even though I can get some python code, it is a black box. So I advocate for as much transparency as possible with these tools - but noting the knowledge gaps that exist. I don't think we should be striving for reproducibility because this is applying a quantitative mindset to qualitative research - I get checking and verifying but 'inter-coder reliability' is a tricky concept in qualitative research so I don't think we should be striving for that. These tools are helpful for reflexivity and making me think about how my interpretations are similar of different to the AI | We appreciate your reflection on transparency limitations and the challenges of reproducibility in qualitative research. Our extension of the COREQ guidelines aims to enhance transparency while acknowledging the inherent complexities and knowledge gaps you identify. We recognize the tension between quantitative notions of reproducibility and qualitative research paradigms, and have designed these guidelines to support reflexivity and methodological transparency rather than imposing rigid reproducibility standards that may be inappropriate for qualitative inquiry. |
| Include spatial LLMs, which integrate language understanding with the analysis of three-dimensional (3D) or spatial data, enabling AI to interpret and reason about physical environments, objects, and their relationships. | Thank you for introducing the consideration of spatial LLMs. We believe the existing reporting items are sufficiently comprehensive to accommodate various LLM architectures, including those that integrate spatial and three-dimensional data analysis capabilities. |
| Thank you very much for sharing this comprehensive survey. I have a few additional suggestions.<br>First, I appreciate the inclusion of reflexivity, but it may also be valuable to integrate researcher positionality into the section on bias. The choice of a particular LLM itself can shape the interpretive process, and acknowledging this positionality could strengthen the guidelines. Relatedly, it may be helpful to ask whether the selected LLM is accessible to stakeholders or participants. While the survey notes whether participants were informed that their data were processed through an LLM, it could also consider participants' digital literacy: many may not fully understand what an LLM is. Even if they do, if the researcher is using a paid or restricted-access model, participants may not be able to engage with it or review outputs during participant validation (a common practice in qualitative research). This issue is particularly important when participants work in different languages or cultural contexts, raising the question of whether the study assessed whether the LLM performed differently across such settings.<br>In addition, given that prompts shape outputs, it may be useful to include a question on whether prompt design was used to bring in a more critical perspective. For example, during analysis, whose voices were amplified, whose were excluded, and were certain perspectives inadvertently silenced? A human coder or checker could play an important role in assessing these dynamics.<br>Finally, I would suggest including an item on conflicts of interest, specifically whether any of the researchers were affiliated with or funded by the developers of the LLMs used in the study. | We appreciate your suggestions. Regarding researcher positionality, we acknowledge that the existing COREQ guidelines already address this fundamental aspect of qualitative research. To specifically capture how researcher positionality influences LLM selection, we have added a new item "Model Selection Rationale" that requires researchers to document their reasoning and positioning in choosing specific models..<br><br>Your points about stakeholder accessibility and participant digital literacy have led us to expand our existing "Informed Consent" item.<br><br>To address your concern about cross-linguistic and cultural contexts, we have implemented a new item "Language considerations" that requires researchers to document the language(s) of interaction.<br><br>Regarding critical prompt design and perspective inclusion, we believe our existing items "User Prompts," "Output Validation," and "Reflexivity" collectively address these concerns. These items require researchers to document their prompting strategies, validate outputs for potential biases or exclusions, and reflect on how their analytical choices, including prompt design, may have amplified certain voices while potentially silencing others.<br><br>Finally, following your recommendation, we have added a dedicated "Conflict of Interest Disclosure" item to ensure transparency regarding any affiliations or funding relationships between researchers and LLM developers. |
| It is important to think through the ethics regarding the use of LLM in analyzing secondary data. For example, do we need to reconsent participants if the de-identified data are used in LLM. | Thank you for highlighting this ethical consideration. We have incorporated this concern by expanded the existing "Informed consent" item to address the need to consider re-consent requirements when using LLMs for secondary data analysis. |
| Suggest to include: statement of possible biases and ways to mitigate them. Explain how sensitive data was protected and whether there were any institutional/regulatory restrictions. Include reliability metrics (e.g. human Kappa vs. LLM) or consensus processes. Report methodological risks associated with the model: biases, limited reproducibility, vendor dependence, etc. Indicate whether prompts, configurations and outputs relevant to traceability were archived | We appreciate your suggestions. We have added the "Interrater-agreement" under the existing item "Output validation" to address reliability metrics including Kappa coefficients. Bias mitigation and methodological risks are covered under the existing "Bias Mitigation" and "Reflexivity" items. The archiving of prompts, configurations, and outputs for traceability is addressed in our documentation requirements. |
| In my opinion, it is better to separate "Don't know" and "Prefer not to answer" as separate response options, as one reflects a lack of knowledge, while the other one reflects a choice to withhold information. In my case, my response would specifically be "Don't know." | Thank you for your methodological observation regarding response options. |
| no suggestions, thanks for such interesting development. Waiting for the final tool. | Thank you for your interest and support. We look forward to sharing the final guideline with the research community. |
| Suggestions for additional reporting items:<br>- Date and time of LLM use<br>- Language of LLM use (and, if applicable, translation of the output)<br>- Technical specifications: e.g., operating system, computer hardware<br>- Confirmation that the approved ethics application of the respective study specifies the use of LLMs | We appreciate your specific suggestions and have incorporated all of them into the guideline: "Session Timestamps" for temporal documentation; "Language considerations" for language specifications and translation processes; "Technical Infrastructure and Environment" for technical specifications; and "Ethical Considerations and Participant Rights" to confirm ethics approval for LLM use. |
| Regarding the following item: "User Prompts"<br>I would suggest including a hint that these details could be included in more depth in the supplementary material, or asking whether these details can be found there. In particular, repeated prompts and revised prompts may make up a long paragraph, which would probably lead to problems with most journals. The same could apply to other items referring to the exact wording of prompts, etc., e.g. the item 'model parameters'. | Thank you for your practical suggestion regarding supplementary materials. We agree that extensive prompt documentation may be better suited for supplementary materials rather than the main manuscript, and this flexibility is built into the guideline implementation. |
| Think of the fact that many users wont have all the skills , It should be easy to use, and yielding quality results | We appreciate your emphasis on usability and accessibility. The guidelines are designed to be implementable by researchers with varying technical expertise while maintaining quality standards. |
| I believe that, in general, the questions are appropriately oriented. However, there are some cases where precision is required for the tool used, which goes against the assumptions of qualitative research. On the other hand, I believe some items related to the interaction between humans and the LLM are missing, especially when the LLM is within and on the circuit. | Thank you for your observation about the balance between precision and qualitative research assumptions. We believe the "Human-LLM Interaction" item adequately addresses the various modes of human-AI collaboration you mention, including situations where the LLM is integrated at different points in the analytical circuit. |
| Why was a certain LLM chosen to the study? For example, I used the OpenAI suite of models because I didn't know I could use the API of others :D | We have added "Model Selection Rationale" to capture the reasoning behind model choices. |
| 1. What language was used or were there translations? 2. Are data protected, confidential and not used to train AI or reproduced without consent/ referencing of authors? | Thank you for these important considerations. We have added "Language considerations" to address language and translation issues, and expanded the existing item "Data privacy and security" to ensure proper data protection and consent protocols are documented. |
| Focusing more on factors such as bias, misinformation, data confidentiality, and non-use by third parties. Revealing an understanding of the Arabic language in context. | We appreciate your emphasis on bias, confidentiality, and linguistic considerations. These concerns are addressed through our new "Prompting lanuage" item for language-specific issues, and revised "Date privacy and security" item. |
| To me there is an importnt balance to be struck - we need guidelines to act more like PRISMA guides for systematic reviews than a tehnical list of tick-boxes.<br>PRISMA helps not just in standardising reporting but also has a vital function in teaching and learning and undertaking a good review by making the key steps explicit and reportable.<br>I therefore feel emphasising credentials sends the wrong message (only skilled operators should be allowed to do this - but who is credintialling them? E.G. as an accredited ATLAS.ti and NVivo and MAXQDA trainer I'm essentially credentialled to act as a salesperson for their software, what is valued by my students is my teaching experience and PhD from a recognised researchn institution. Those credentials say much less.)<br>Therefore I feel that details to allow reproducibility (name of model, example of prompts) are good for "doing science", however fundamental qualitative considerations and accounting for those through detailing steps of reflexivity, bias, consent and ethics are ***absoltuely key considerations***. These serve not only to increase trustworthiness but also guide new users on considerations of intentional, critical use and therefore not only help in learning and teaching but also should help encourage and strcuture reflection by expereinced users. | We appreciate your comparison to PRISMA guidelines and your emphasis on the pedagogical function of reporting guidelines. We agree that credentials should not create barriers to entry but rather promote transparency. The guideline prioritizes fundamental qualitative considerations including reflexivity, bias, consent, and ethics as essential elements. These items serve both to enhance trustworthiness and to guide researchers in developing critical and intentional approaches to LLM integration in qualitative research. |

| | |
|---|---|
| The importance of a lot of these items would depend on the way in which LLMs were being used in qualitative analysis. For example, time efficiency would be very important if you are developing a tool to analyse qual data, but not if you are using an LLM to just gather insights. Some also depend a lot on if you are running an LLM via an API or using a chat window - so perhaps in the guidelines this should be distinguished more.<br><br>Also, I think the bias mitigation item should be separated into 1) biased training data, and 2) triangulation with other methods, as these feel like quite different items. | Thank you for your observations. We have added "Model Selection Rationale" to help researchers explain their specific use cases and implementation choices. Regarding bias mitigation, we believe the current structure with "Reflexivity" addressing training data concerns and "Output Validation" covering triangulation provides appropriate coverage while maintaining clarity. |
| The hosting environment for the LLM. E.g. commercial provider (if so which one), locally hosted on device, institutional server | We appreciate your suggestion regarding hosting environment documentation. This has been incorporated into the new item "Technical Infrastructure and Environment". |
| No, it seems to be a comprehensive list. | Thank you for your feedback. |
